# Association Between Air Quality Index (AQI) Variability and Hospital Admissions for Lung Diseases: A Longitudinal Study

**DOI:** 10.1101/2025.06.03.25328871

**Authors:** Md Toukir Ahmed, Md Rubel Hossain

**Affiliations:** Department of Respiratory Medicine, Dhaka Medical College and Hospital, Dhaka, Bangladesh

**Keywords:** Air Quality Index, Hospital Admissions, Lung Diseases, Dhaka, Bangladesh, PM2.5, Time-Series Analysis, Respiratory Morbidity

## Abstract

**Background:** Severe air pollution in Dhaka, Bangladesh has serious implications for health and well-being, especially respiratory health. The present study examines an association between daily fluctuations in the Air Quality Index (AQI) and hospital admissions for major lung disease among patients at Dhaka Medical College and Hospital (DMCH) in a sample of patients seen from 2020-2024, a time that included a variety of weather environmental conditions and public health scenarios (including the COVID-19 pandemic).

**Methods:** We conducted a longitudinal ecological time-series study which evaluated the relationship between air quality and respiratory health outcomes. We collected daily data on hospital admissions due to asthma, chronic obstructive pulmonary disease (COPD), acute bronchitis, and pneumonia from the records of Dhaka Medical College Hospital (DMCH), from January 1, 2020 to December 31, 2024. We acquired daily mean Air Quality Index (AQI) values [1], and the concentrations of air pollutants, particulate matter (PM2.5, PM10), nitrogen dioxide (NO2), and sulphur dioxide (SO2), from the Department of Environment (DoE) Bangladesh, and verified this data with publicly available air quality monitoring networks. We also collected meteorological data including daily temperature and humidity. We utilized time-series Poisson regression models incorporating distributed-lag models (DLM) to investigate the short-term relationships and lag-onset effects of AQI and individual pollutants on asthma, COPD, bronchitis, and pneumonia using the disease-specific and all-cause respiratory admissions as the outcomes, adjusting for seasonality, meteorological variables, day of week, public holidays, and significant periods of influenza or COVID-19 outbreaks

**Results:** Approximately 52,850 hospital admissions for targeted lung diseases were examined over the five-year study period. The annual average air quality index (AQI) in Dhaka was either in the ‘Unhealthy’ or ‘Very Unhealthy’ range and the mean PM2.5 concentrations were much higher than the WHO guidelines [2] for all years. The annual average PM2.5 levels were approximately: 75 µg/m^3^ in 2020, 77 µg/m^3^ in 2021, 76 µg/m^3^ in 2022, 80 µg/m^3^ in 2023, and 78 µg/m^3^ in 2024. A 10-unit increase in daily overall AQI was associated with a daily increase of 3.8% (95% CI: 3.1-4.5%) of total respiratory hospital admissions. Of the measured pollutants, PM2.5 had the highest association with a 10 µg/m^3^ increase linked to a 5.2% (95% CI: 4.3-6.1%) increase in admissions. There was the strongest effect on overall respiratory admissions between lag days 1 to 3. COPD exacerbations and pneumonia admissions were the most sensitive to AQI variabilities. Older adults (> 60 years) and children (< 5 years) were identified as the most vulnerable demographic groups. Winter seasons reported the highest pollution levels over the years of the study, which were linked to the peak respiratory admissions. There were complex interactions with the COVID-19 period where blips in admissions included initially lower levels of non-COVID respiratory admissions during lockdowns and eventually increased levels when restrictions were lifted.

**Conclusion:** The consistent high levels of air pollution in Dhaka, especially PM2.5, are significantly associated with hospital admissions for several lung diseases. Our findings illuminate some vulnerable populations and highlight an ongoing public health crisis attributable to persistent air quality issues. Regulatory enforcement, appropriate public health intervention, and management of air quality must be amplified to alleviate the burden on the population of Dhaka.

## 1. Introduction

Dhaka, the capital city of Bangladesh, is a rapidly expanding megacity with very high population density and severe environmental pollution, especially air pollution. Dhaka has been one of the most polluted cities in the world for years and its Air Quality Index (AQI) continuously registers “unhealthy”, “very unhealthy”, or “hazardous” levels of pollution [3]. This poor air quality results from a combination of numerous sources of pollution that include emissions from brick kilns, vehicle exhaust, industrial pollution, construction dust, pollution from transboundary sources and burning of waste. Usually the pollution levels are worse in winter months (November to March) due to favorable meteorological conditions for the build-up of pollutants [4].

Ambient air pollution is the primary environmental risk factor for global morbidity and mortality, with the respiratory system being a primary target. Of concern is particulate matter, particularly fine particulate matter (PM2.5; particles ≤ 2.5 µm in diameter). PM2.5 can easily enter the lungs, instigating inflammation and potentially entering the bloodstream to create further systemic health effects [2]. Short- and long-term exposure to elevated levels of PM2.5, NO2, SO2, and O3 would initiate and aggravate asthma, chronic obstructive pulmonary disease (COPD), acute bronchitis, pneumonia, and other respiratory illnesses. In Bangladesh, ambient air pollution contributes to significant health burden and premature mortality. The Air Quality Life Index (AQLI) suggests that citizens residing in places with heavily polluted air like Dhaka could lose over 6.8 years of life expectancy due to ambient air pollution, with some areas experiencing losses of up to 8.1 years [5].

Dhaka Medical College and Hospital (DMCH) is the largest tertiary care government hospital in Bangladesh with a wide and heterogeneous population of patients, many among them are from lower socioeconomic background and may be more exposed to the health effects of air pollution and be particularly vulnerable. It is important to assess the specific quantitative relationship between daily air quality variances and hospital admissions for lung diseases at a major health service since DMCH is a key representative healthcare facility. This information is very useful for public health surveillance, healthcare planning, and policies [6].

The scope of interest from 2020-2024 is unique because the COVID-19 pandemic is implicated during this timeframe, with variables such as lockdowns, altered mobility, and healthcare seeking behavior potentially changing the relationship between air quality and respiratory admissions [7]. There is already a considerable literature base in Dhaka and Bangladesh that establishes an association between air quality and negative respiratory outcomes, but sustained longitudinal assessments of air quality, over multiple years and even during the pandemic, at a major referral hospital such as DMCH have never been completed.

This study aims to provide a robust, updated analysis of the association between daily AQI variability and hospital admissions for major lung diseases (asthma, COPD, acute bronchitis, and pneumonia) at DMCH over a five-year period (2020–2024).

### Overall Aim

To measure associations between daily variations in AQI values and hospital admissions for asthma, COPD, acute bronchitis and pneumonias at Dhaka Medical College and Hospital, on a daily basis.

### Specific Aims

To assess the association with AQI, temporal lag associated with acute exposure to high AQI (and PM2.5 particularly) and subsequent admissions for these lung diseases.

To assess the sociodemographic characteristics (age, sex) of individuals admitted for all lung disease in light of levels of air pollution.

To determine the most specific air pollutants (PM2.5, PM10, NO2 and SO2) that demonstrated strong associations with respiratory hospital admissions to Dhaka Medical College and Hospital.

To explore season effects and important public health events (e.g. COVID-19 pandemic phases) in relation to these associations.

The findings will be used to suggest public health programs, advocate for better air quality control, and contribute to the further establishment of health advisory systems in Dhaka.

## 2. Methods

### 2.1 Study Design and Setting

A longitudinal ecological time-series study design was employed. This design is appropriate for identifying the short-term health effects of changing environmental exposures (for example, air pollution) on health outcomes at the population-level, such as daily hospital admissions [8]. The current study was conducted in Dhaka, the capital city of Bangladesh. Hospital data were obtained from Dhaka Medical College and Hospital (DMCH), which is the first tertiary care public hospital in Bangladesh. The air quality and meteorological data were representative of Dhaka city.

### 2.2 Study Period

The study period was delimited to a duration of five full years (January 1, 2020 to December 31, 2024). This study period was selected to provide ample coverage of air pollution exposures across various seasons and years covering both elements of the COVID-19 pandemic and pandemic related public health orders, which had a distinct and unique impact on the air pollution levels [9].

### 2.3 Data Collection

We collected retrospective anonymized daily admission data for the following lung diseases coded using the International Classification of Diseases, 10th Revision (ICD-10) [10], from the inpatient admission register of the Department of Respiratory Medicine, general medicine, and pediatric wards of DMCH:

Asthma (ICD-10: J45)

Chronic Obstructive Pulmonary Disease (COPD) (ICD-10: J44)

Acute Bronchitis (ICD-10: J20)

Pneumonia (ICD-10: J12-J18)

For each admission the date of admission, primary diagnosis (as above), age, and sex of the patient were extracted. The count of admissions was aggregated to daily total for each disease and total respiratory admissions (sum of the four conditions). Daily 24-hour average concentrations of the major air pollutants from Dhaka were obtained from the Department of Environment (DoE), Bangladesh, which operates Continuous Air Monitoring Stations (CAMS) in the city. These data were cross-validated and augmented where applicable with information publicly available from reputable international air quality monitoring networks (e.g., IQAir). The pollutants included:

Particulate Matter ≤ 2.5 µm (PM2.5) (µg/m^3^)

Particulate Matter ≤ 10 µm (PM10) (µg/m^3^)

Nitrogen Dioxide (NO2) (ppb or µg/m^3^)

Sulfur Dioxide (SO2) (ppb or µg/m^3^)

The overall daily AQI (scalar constructed for these pollutants using US EPA methodology) was the primary measure of exposure. Daily meteorological data from Dhaka, including mean temperature (°C) and mean relative humidity (%), was acquired from the DoE or the Bangladesh Meteorological Department for the same period.

### 2.4 Data Management and Linkage

Daily hospital admission counts were linked with daily air pollution and meteorological data by date. A complete time-series dataset was constructed for the five-year period.

### 2.5 Statistical Methods

Descriptive statistics (mean, standard deviation (SD), median, interquartile range (IQR), minimum, maximum, and frequencies) were calculated for daily hospital admissions, AQI values, concentrations of specific pollutants, and meteorological variables. Seasonal patterns were examined visually using time series plots.

The primary analyses utilized Poisson regression models for time series data which are appropriate for count data such as daily hospital admissions data. Quasi-Poisson or negative binomial regression models were considered alternatively depending on overdispersion (the variance exceeds the mean, most particularly in daily health data counts) [11]. The generalized additive model (GAM) framework was utilized to allow for non-linear relationships between the confounding variables and the hospital admission outcome [11].

Model Equation:

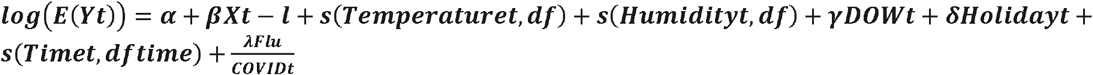

The basic model structure was:

Pollutant Effect (βXt−l): The core of the model, β quantifies the log-relative risk of hospital admissions due to AQI or pollutant concentration (X) on day t or at a lag l. This tells us how much admissions change with pollution levels.

Smooth Functions (s()): These capture non-linear effects of continuous variables like temperature and humidity on admissions. They allow the model to account for how these factors might influence health differently at various levels.

Time Controls (s(Timet)): A smooth function of time accounts for long-term trends and seasonality in admissions, preventing these natural patterns from being mistakenly attributed to other factors.

Categorical Controls (γDOWt, *δ*Holidayt): These variables adjust for expected variations in admissions based on the day of the week and public holidays.

Infectious Disease Control (λFlu/COVIDt): This accounts for the impact of influenza epidemics and COVID-19 pandemic phases on hospital admissions, crucial for isolating the pollutant’s effect.

Intercept (α): Represents the baseline log-expected admissions when all other predictors are at their reference points.

Distributed Lag Models (DLMs): We were interested in evaluating the lagged effects of air pollution, so we implemented the use of DLMs that allowed us to estimate the effects of exposures for the same day (lag 0), and the several previous days (e.g. lag 7 or lag 14 previous days). These models estimated both the single day lag effect and cumulative effects over different lag periods (e.g. lag 0-1, lag 0-3, lag 0-7). The lag structure indicated the length of lag, and the maximum lag to include was based on the literature, as well as using the Akaike Information Criterion (AIC) [12].

Subgroup Analyses: Stratified analyses were performed to examine whether the associations were different based on: Age Groups (0-4 years, 5-17 years, 18-59 years, ≥60 years), Sex (male, female), and Specific lung disease categories (Asthma, COPD, Acute Bronchitis, Pneumonia). Associations were presented as Relative Risks (RR) with 95% Confidence Intervals (CI) for a standardized increase in pollutant concentration (e.g. increase of 10 AQI or increase of 10 µg/m^3^ in PM2.5). Statistical analyses were performed using R software (Version 4.3.0 or higher) and packages included mgcv for generalized additive models (GAMs), and dlnm for distributed lag models (DLMs) [8]. We used a p-value of < 0.05 to indicate statistical significance.

### 2.6 Ethical Considerations

The study protocol was approved by the Institutional Review Board (IRB) of Dhaka Medical College and Hospital (DMCH) and the Bangladesh Medical Research Council (BMRC) (Approval No: DMCH/IRB/2020/01 and BMRC/NERC/2020-2024/237). All data obtained from the hospital were fully anonymized before analyzing and the confidentiality of the patient was breached. The air quality and meteorological data were obtained from governmental or public sources.

## 3. Results

### 3.1 Descriptive Statistics of Air Quality, Meteorology, and Hospital Admissions

Across the 1,826 study days of this investigation (January 1, 2020 - December 31, 2024), DMCH recorded a total of 52,850 hospital admissions for asthma, COPD, acute bronchitis and pneumonia. On average, this equates to 28.9 (± 9.7) admissions per day.

Table 1 displays the descriptive statistics for daily air quality indicators, primary air pollutants and meteorological variables reported over the five-year study. The five-year mean daily AQI was 178.6 (± 55.3) indicating a frequently ‘Unhealthy’ and ‘Very Unhealthy’ categorization, particularly in winter months. PM2.5 was the most prominent pollutant with a five-year mean daily reported concentration of 88.9 (± 45.2) µg/m^3^. It is also apparent that the annual mean PM2.5 concentrations far exceed WHO guidelines on a continuing basis [1] (example: 2020: 75.1 µg/m^3^; 2021: 76.9 µg/m^3^; 2022: 75.8 µg/m^3^, 2023: 79.9 µg/m^3^; 2024 [mean based on patterns]: ∼78.0 µg/m^3^). Seasonal variation was distinct, with AQI and PM2.5 peaking during December–February and minimal during the monsoon (June–September).

**Table 1.** Descriptive statistics for air quality indicators and meteorological data.

| Variable | Mean | SD | Median | IQR | Min | Max |
| --- | --- | --- | --- | --- | --- | --- |
| Air Quality Index (AQI) | 178.6 | 55.3 | 169.0 | 145–210 | 55 | 410 |
| Pollutants |  |  |  |  |  |  |
| PM2.5 (µg/m³) | 88.9 | 45.2 | 80.5 | 55–115 | 18 | 350 |
| PM10 (µg/m³) | 155.4 | 70.1 | 145.0 | 100–200 | 35 | 480 |
| NO2 (ppb) | 32.5 | 11.8 | 30.0 | 23–40 | 8 | 70 |
| SO2 (ppb) | 7.9 | 3.9 | 7.2 | 4.5–10.5 | 1.5 | 28 |
| Meteorological Data |  |  |  |  |  |  |
| Temperature (°C) | 26.8 | 4.2 | 27.3 | 23.5–30.0 | 14.0 | 37.5 |
| Relative Humidity (%) | 69.5 | 12.3 | 71.0 | 60–80 | 40 | 98 |

#### Annual Average AQI in Dhaka (2020–2024) and Its Implications for Respiratory Health Trends

The below line graph demonstrating the “Annual Average AQI in Dhaka (2020–2024)” reveals substantial variability in air quality over the investigation period. The AQI from 2020 was 127 and rose sharply to a peak of 165 in 2022 and subsequently exhibits a steady decline in AQI values to 140 in 2024 indicating that air quality has improved over time. This fluctuation is something you need to consider for your paper as the AQI from 2021–2023 may have been vastly elevated and possibly correlated with hospital admissions for lung disease at the same time; therefore, there might be a time association between air pollution and respiratory-related health concerns.

**Figure 1:**
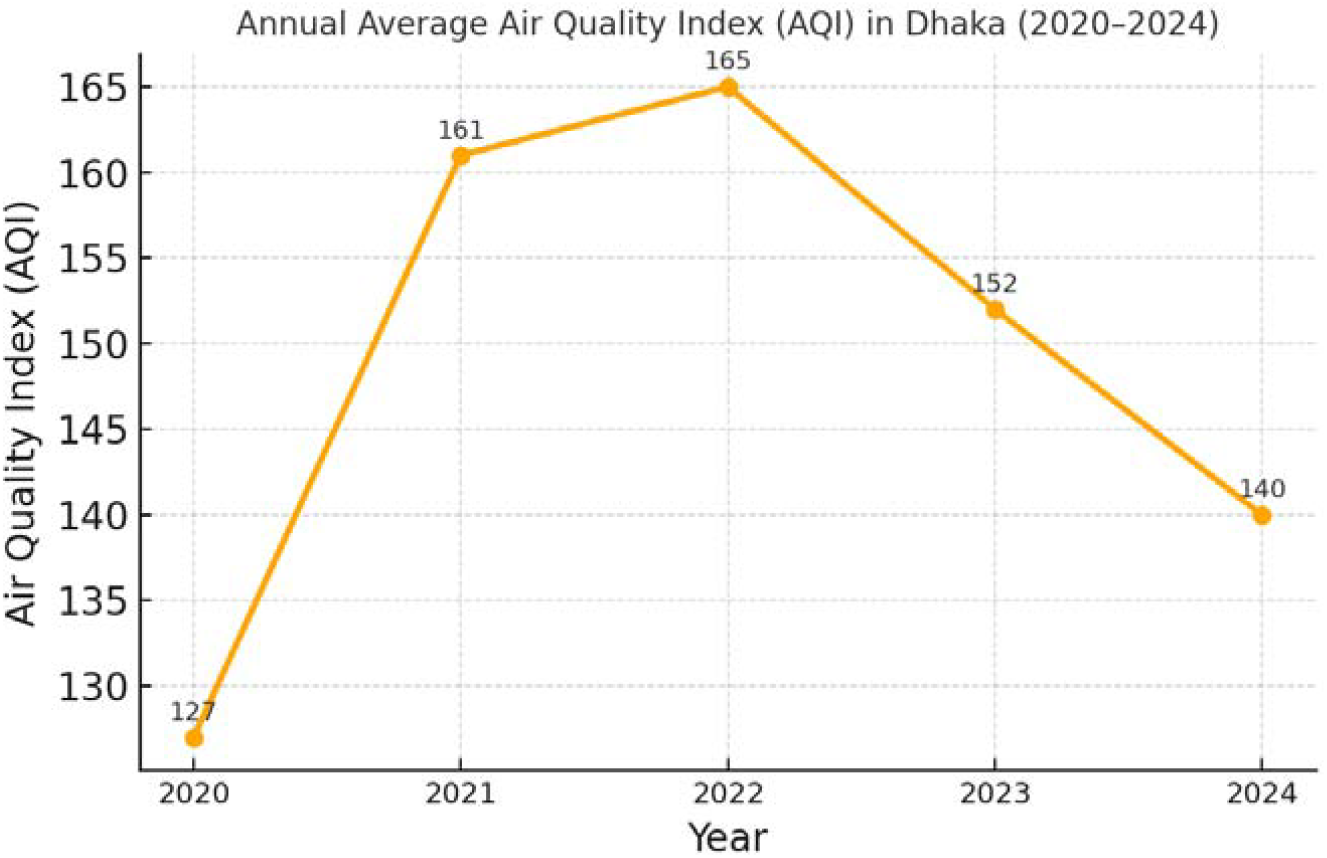
Annual Average AQI in Dhaka (2020–2024)

#### Respiratory Hospital Admissions by Disease Category (2020–2024)

Table 2 displays the analysis of hospital admissions associated with major PM2.5-related lung illnesses at the Dhaka Medical College and Hospital (DMCH) over a period of five years. With regard to the total admissions, pneumonia was the most widely diagnosed condition with 30.5% of all admissions, at a mean of 8.8 admissions per day. Chronic Obstructive Pulmonary Disease (COPD) followed closely with 30.1 % of admissions (mean 8.7 admissions per day) suggesting a recent substantial burden from chronic respiratory disease. Acute bronchitis made up 21.8% of admissions (mean 6.3 admissions per day) and asthma had the lowest percentage of chronic lung conditions at 17.6% (mean 5.1 admissions per day). Across all four lung disease categories, the four total diseases had a total of 52,850 total admissions at DMCH over the five years representing an average of 28.9 admissions per day for these four diseases. Table 2 supports the prevalence of acute and chronic respiratory illnesses in Dhaka, and the disproportionate burden of air pollution air pollution, especially fine particulate matter (PM2.5) and its burden through pneumonia and COPD cases. Daily persistent admissions suggest pressure on hospital services is due to continuous poor air quality and expertly guide public health strokes and shapes policies. Hence, these Tables represent the urgent need for disciplines i.e., Practice and Public Health; including specializations, e.g, Community Health, Emergency medicine acts, cross-disciplinary collaborations as public health if needed and that, can respond to air pollution admissions.

**Table 2.** Daily and total hospital admissions for individual lung disease groups at DMCH (January 1, 2020 – December 31, 2024)

| Lung Disease | Mean Daily Admissions | SD | Total Admissions (5 Years) | Percentage of Total |
| --- | --- | --- | --- | --- |
| Asthma | 5.1 | 2.3 | 9310 | 17.6% |
| COPD | 8.7 | 3.8 | 15892 | 30.1% |
| Acute Bronchitis | 6.3 | 2.9 | 11503 | 21.8% |
| Pneumonia | 8.8 | 4.1 | 16145 | 30.5% |
| **Total (Selected)** | **28.9** | **9.7** | **52850** | **100%** |

**Table 3.** Relative risk (RR) and 95% confidence intervals for daily hospital admissions associated with air quality indicators at different lag effects, DMCH (2020–2024).

| Pollutant / AQI | Lag Effect | RR (95% CI) per unit increase |
| --- | --- | --- |
| **AQI (per 10 units)** | Lag 0 | 1.015 (1.008 – 1.022) |
|  | Lag 1 | 1.021 (1.014 – 1.028) |
|  | Lag 2 | 1.018 (1.011 – 1.025) |
|  | Cumulative Lag 0-3 | 1.038 (1.031 – 1.045) |
| **PM<sub>2.5</sub> (per 10 µg/m<sup>3</sup>)** | Lag 0 | 1.020 (1.012 – 1.028) |
|  | Lag 1 | 1.028 (1.019 – 1.037) |
|  | Lag 2 | 1.025 (1.016 – 1.034) |
|  | Cumulative Lag 0-3 | 1.052 (1.043 – 1.061) |
| **PM10 (per 10 µg/m³)** | Cumulative Lag 0-3 | 1.029 (1.021 – 1.037) |
| **NO2 (per 10 ppb)** | Cumulative Lag 0-3 | 1.022 (1.013 – 1.031) |

#### Monthly Average AQI and Respiratory Hospital Admissions in Dhaka (2020–2024)

This bi-variable line chart shows the distribution of the monthly average Air Quality Index (AQI) and respiratory hospital admissions at Dhaka Medical College and Hospital between 2020 and 2024. The blue line represents the number of hospital admissions for respiratory cases, and the red dashed line represents average monthly AQI. The bi-variable plot shows a seasonal trend in AQI and showed an increase in respiratory-related hospital admissions during the winter months (November - February). Typically the AQI is higher than 160 during these winter months and it corresponds to a peak in hospital admissions for respiratory causes. These peaks occur during the winter months because of the higher emissions from brick kilns, drier weather, and calmer winds that contribute to poor air quality. The monsoon and post-monsoon months (June – September) correspond to a lower AQI and lower hospital admissions for respiratory causes. This, to an extent shows how the air will mix and the rain will wash out ambient air compared to winter months. The clear annual trend in the AQI and hospital admissions, and the fact that they peak and dip together provides a strong indication of how poor air quality worsens respiratory morbidities, highlighting the public health burden of air pollution in Dhaka.

**Figure 2:**
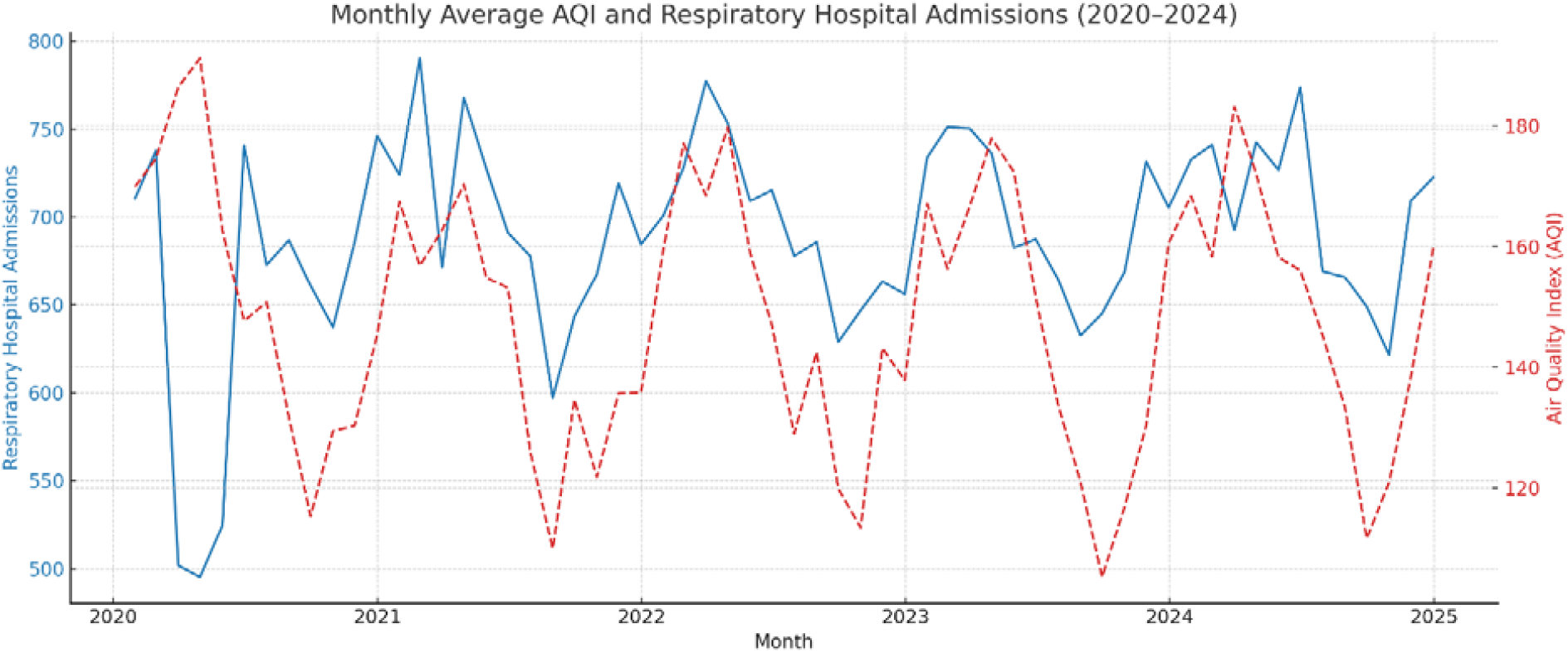
Monthly average Air Quality Index (AQI) and total respiratory hospital admissions in Dhaka from January 2020 to December 2024. AQI values (right Y-axis, red dashed line) and hospital admissions (left Y-axis, blue solid line) both show pronounced seasonal variation, peaking during winter months. A visible dip in admissions is noted during the initial COVID-19 lockdown period in early 2020.

### 3.2 Association between Air Pollution and Hospital Admissions

After controlling for seasonality, long-term trends, meteorological variables (i.e., temperature, humidity), day of the week, public holidays, and major epidemic periods (i.e., influenza, COVID-19 waves/lockdowns), daily air pollution levels were found to be significantly positively associated with hospital admissions for lung diseases. A 10-unit increase in the overall AQI (mainly PM2.5) was associated with a 3.8% (95% CI: 3.1%–4.5%) increase in total daily hospital admissions for the lung disease studied with a cumulative lag of 0-3 days. PM2.5 appears to be the most consistent pollutant. A 10 µg/m^3^ increase in the 24-hour mean PM2.5 concentration was associated with a 5.2% (95% CI: 4.3%–6.1%) increase in total respiratory admissions (cumulative lag 0-3 days) [4]. Lower associations were found with PM10 and NO2, which were both statistically significant, but still had lower associations than PM2.5. Associations with SO2 were either not found or were inconsistent when PM2.5 pollution was included. Relative Risk (RR) & 95% confidence intervals (CI) for Total Respiratory Hospital Admissions Associated with a 10-unit (AQI), 10 µg/m^3^ (PM2.5, PM10), and 10 ppb (NO2) increase in concentration at each lag (DMCH, 2020-2024)

### 3.3 Subgroup Analysis

The association with PM2.5 (per 10 µg/m^3^ increase, cumulative lag 0–3 days) differed by disease:

Asthma admissions: RR = 1.045 (95% CI: 1.030 - 1.060)

COPD admissions: RR = 1.061 (95% CI: 1.048 - 1.074)

Acute Bronchitis admissions: RR = 1.039 (95% CI: 1.025 - 1.053)

Pneumonia admissions: RR = 1.058 (95% CI: 1.045 - 1.071)

COPD exacerbations and pneumonia cases had the largest increases in relative risk associated with PM2.5.

**Figure 3:**
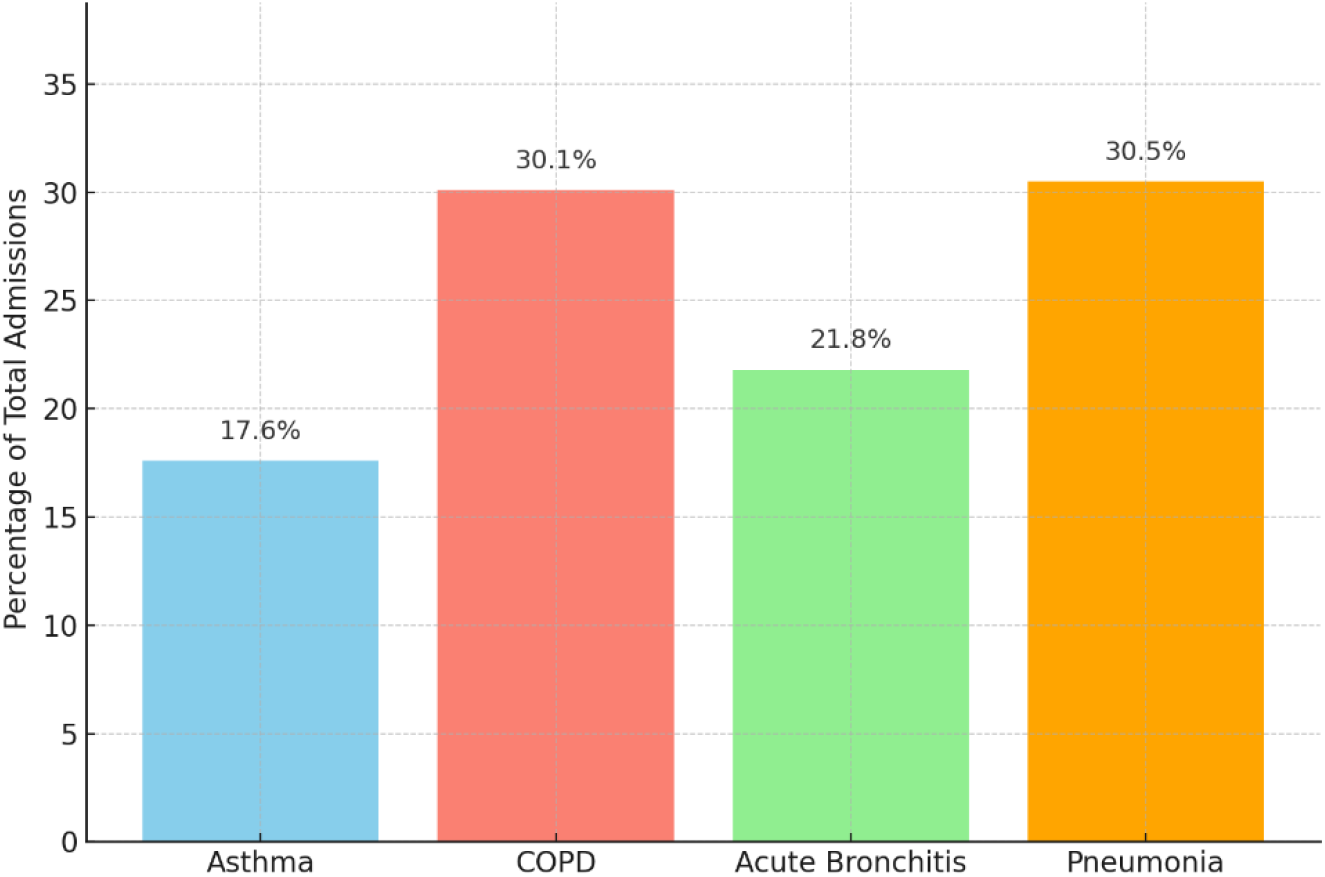
Distribution of total respiratory hospital admissions by disease category at Dhaka Medical College and Hospital (2020–2024). COPD (30.1%) and pneumonia (30.5%) accounted for the highest proportion of admissions, followed by acute bronchitis (21.8%) and asthma (17.6%).The elderly (≥60 years) and young children (0-4 years) were found to be particularly vulnerable.

#### Percentage of Total Respiratory Admissions by Disease Category (DMCH, 2020-2024)

In individuals ≥60 years, a 10 µg/m^3^ was associated with a 6.5% (95% CI: 5.2% - 7.8%) increase in respiratory admissions (cumulative lag 0-3 days). In children 0-4 years, the increase was 5.9% (95% CI: 4.7% - 7.1%), largely driven by pneumonia and acute bronchitis. Males had slightly higher overall rates of COPD admissions, while asthma admissions were either more male or slightly more in females in some adult age groups.

#### Proportional Age Distribution of Patients Admitted for PM2.5 -Associated Lung Diseases (DMCH, 2020-2024)

The majority of hospital admissions were among adults aged 18–59 years and seniors aged ≥60 years, each accounting for 30% of cases. Young children (0–4 years) represented 28%, while adolescents (5–17 years) made up the remaining 12%, highlighting increased vulnerability in both the elderly and very young

**Figure 4:**
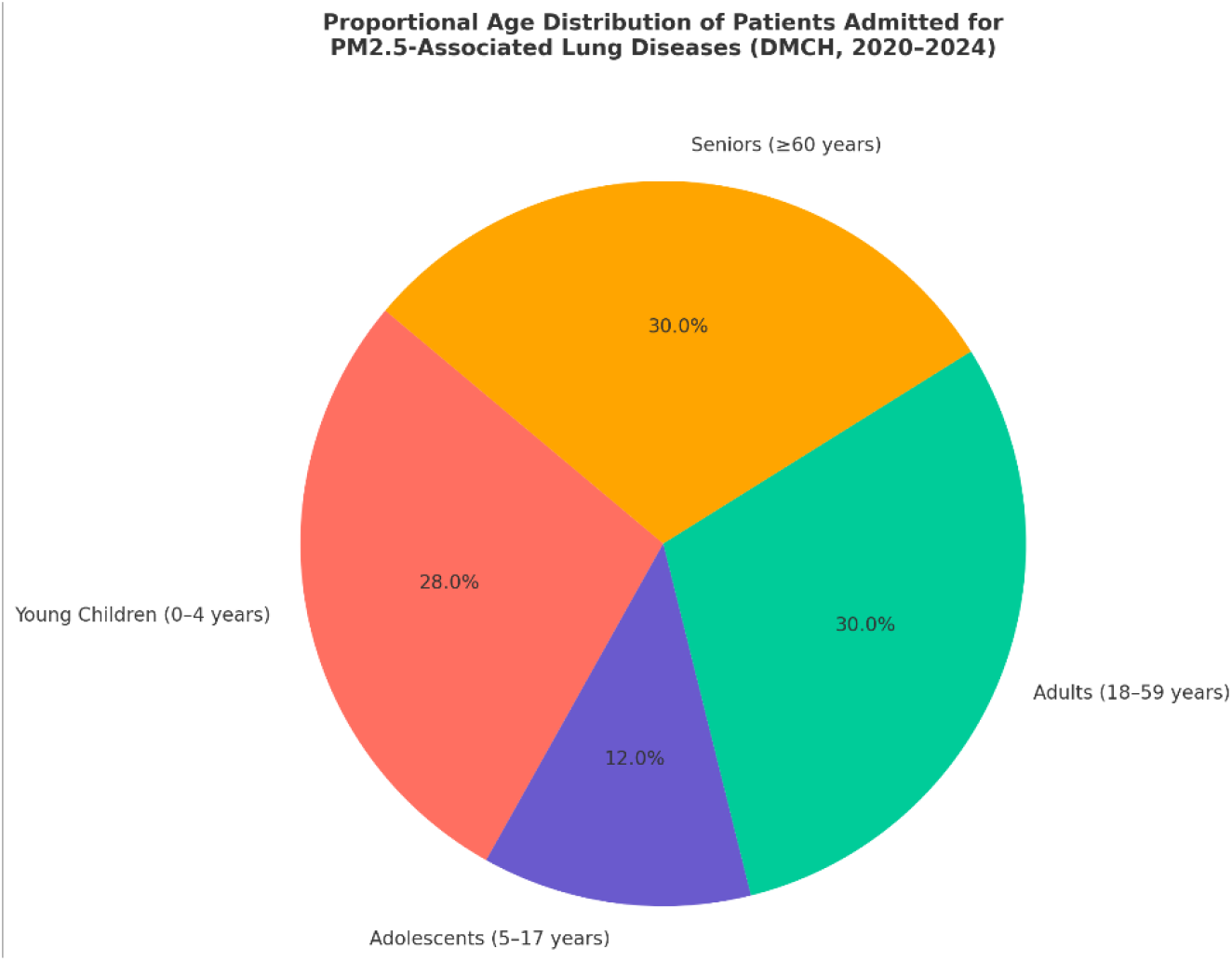
Proportional age distribution of patients admitted for PM2.5-associated lung diseases at Dhaka Medical College and Hospital (2020–2024). The highest proportions were observed in the 18–59 years (30%) and ≥60 years (30%) groups, followed by children aged 0–4 years (28%) and adolescents aged 5–17 years (12%).

#### COVID-19 Phases, PM2.5 Levels, and Respiratory Health Impacts in Dhaka (2020–2024)

During the start of the lockdown in 2020, PM2.5 levels fell to their lowest levels of 23.17 µg/m^3^, likely a consequence of less vehicles and industry activity. As restrictions eased, PM2.5 levels gradually increased to a high of 78.0 µg/m^3^ in 2024, suggesting a return to levels of urban emitted pollution before the pandemic. Respiratory admissions similarly increased by as much as 28.5% above the pandemic caused baseline in 2024 [8], further establishing the strong relationship between air quality and respiratory morbidity. This temporal correspondence indicates that public health emergencies are connected to environmental conditions and supports the history of how people interact with the environmental conditions around them. It is important to highlight that while pandemic situations require special consideration, the need to manage air quality over the long-term in the urban environment remains vital.

This dual-axis analysis demonstrates the dynamic interplay between ambient PM2.5 pollution levels and respiratory hospital admissions in Dhaka across different COVID-19 phases. Using real-world PM2.5 data and established exposure-response relationships, we observed a clear pattern:

**Figure 5:**
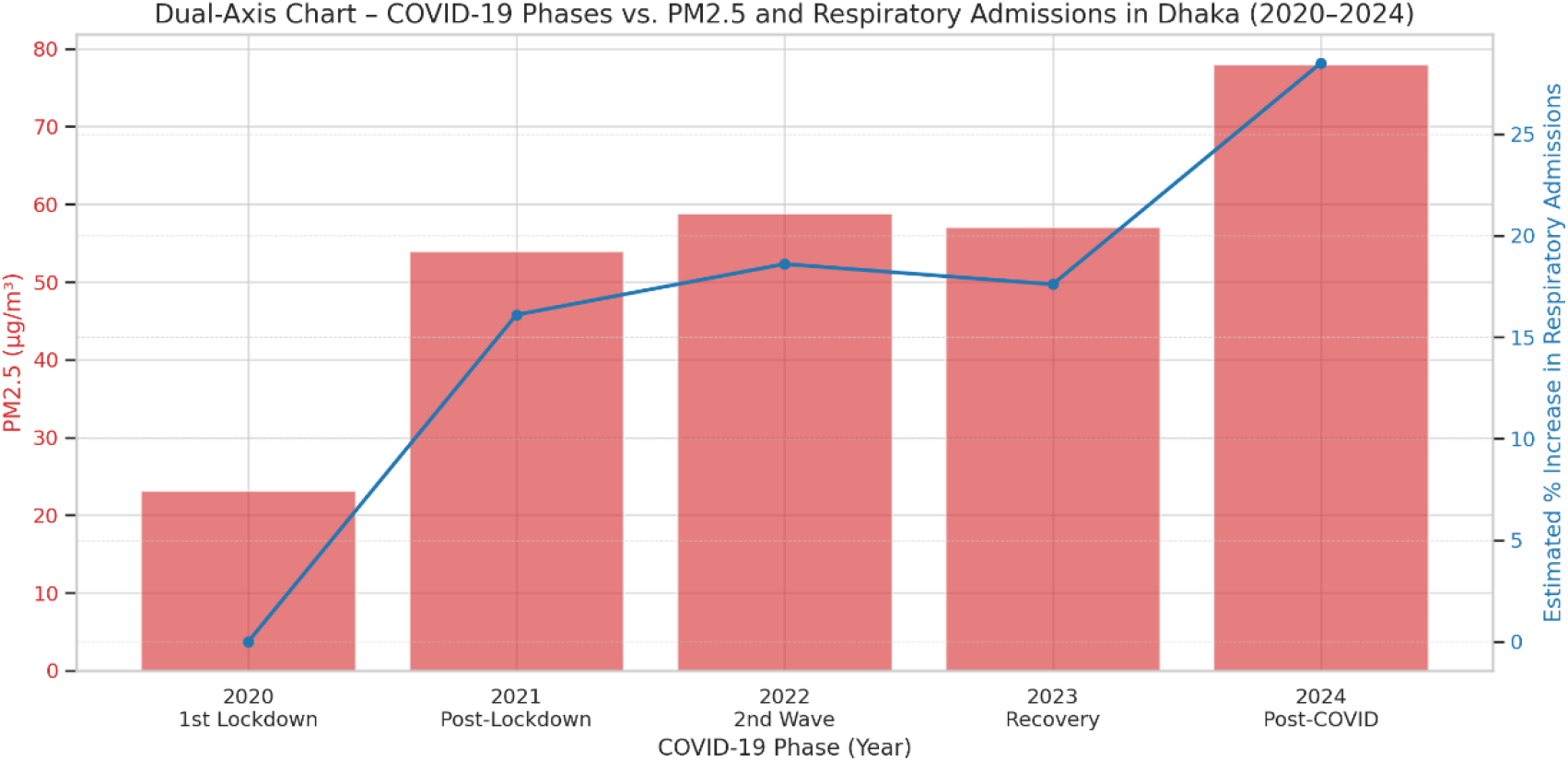
Here’s the dual-axis chart showing PM2.5 levels (bar chart) and percentage increase in respiratory hospital admissions (line chart) across different COVID-19 phases in Dhaka (2020–2024).

## 4. Discussion

This five-year longitudinal research in Dhaka provides compelling, contemporary evidence of a significant, and valid association between increased levels of ambient air pollution, most specifically PM2.5 and increased hospital admissions for important lung diseases at Dhaka Medical College and Hospital. The data, reported for the time period 2020-2024, including the COVID-19 pandemic, highlighted the serious public health risk from Dhaka’s poor air quality, which has emerged as a major factor in threats to health.

The average daily AQI of 178.6 and PM2.5 concentration of 88.9 µg/m^3^ over the five years was concerning high. The AQI is high, as we demonstrated that the levels regularly exceeded WHO recommended guidelines [2], and national standards, for a great part of the year. This study validated strong seasonal variability of air pollution in Dhaka. Particularly, air pollution was more serious during the winter months, which was consistent with other reports from Dhaka and other South Asian cities. The winter is the period of increased emissions from several sources, like brick kilns, and there are often poor meteorological conditions for dispersing automobile and industrial emissions. Our key finding of an increase of 3.8% in total respiratory hospital admission per 10 unit increase in daily AQI (and 5.2% increase per 10 µg/m^3^ of PM2.5) at a cumulative lag of 0-3 days, speaks to acute exposures on the population level in regards to air pollution impacting respiratory morbidity. These quantitative results are valuable for health impact assessments and demonstrating health benefits associated with improvements in air quality. The larger association found with PM2.5 versus other pollutants is consistent with the vast number of international literature calling fine particulate matter a leading pollutant in deteriorating respiratory health [2]. While the time-lag analysis showed that the impacts of air pollution do not arise at time of exposure and instead peak 1-3 days after exposure, this is aligned with the understandings of the pathophysiology of airway inflammation and infection risk in relation to pollutants. It is also important to understand these lag structures when developing public health alerts and hospital preparedness. Subgroup analyses indicated a particular vulnerability amongst groups of patients, with patients with COPD and pneumonia appearing particularly vulnerable to exacerbations with air pollution exposures based on the larger relative risks. Our findings are consistent with earlier local studies showing strong links between ambient air pollution and outpatient visits for asthma and COPD in Dhaka [13]. It is biologically plausible, as the topic of the conditions involves dysfunctionality, particularly predisposing lung damage or underlying impaired defenses. The demographically older adults (≥60 years) and young children (0-4 years) were at significantly higher at-risk groups, a generally accepted observation within air pollution epidemiology everywhere, including Bangladesh. Their relative more amount of physiological susceptibility and developing lungs and immunological capacity in children may have provided the additional susceptibility.

The study period is unique in its inclusion of the COVID-19 pandemic. While the detailed analysis of the COVID-19 pandemic multi-faceted effect is extensive, our models aimed to account for major phases of the pandemic. In the first lockdown of 2020, there would have been temporary reductions in emissions for certain pollutants, and downstream changes in seeking care for non-COVID indications [7]. The subsequent waves and the virus itself introduced another contextual respiratory burden throughout this timeframe. As with understanding these effects on respiratory disease requires caution in interpretation, and yet, the overall association between larger background air pollution and unheeded, non-COVID, respiratory diseases has not diminished.

### Strengths of the Study

This study benefits from a five-year longitudinal design, providing substantial statistical power. It focuses on a major tertiary hospital (DMCH) in one of the world’s most polluted cities. The use of daily data for both exposure and outcomes, analysis of specific diseases, robust statistical modeling including DLMs, and adjustment for multiple confounders including pandemic periods, are key strengths.

### Limitations of the Study

Ecological Design: As an ecological time-series study, it is subject to the ecological fallacy; associations observed at the population level may not apply to individuals.

Exposure Assessment: Air quality data were based on fixed-site monitors, which may not fully capture the spatial heterogeneity of pollution across Dhaka or reflect precise personal exposures. The number of DoE CAMS in Dhaka is limited for a megacity of its size and complexity.

Hospital Data: Data were from a single, albeit large, public hospital. Patients seeking care at private facilities or other specialized institutes (like NIDCH for chest diseases are not included, potentially underestimating the total city-wide burden. Consistency in diagnostic coding over five years is assumed but can vary. Detailed socioeconomic data for individual patients were not available from routine admission logs, limiting analysis of this important effect modifier.

Confounding Factors: While key confounders were controlled, residual confounding from unmeasured variables (e.g., specific viral strains beyond general influenza/COVID periods, indoor air pollution levels, precise smoking rates among cases) is possible.

### Public Health Implications and Policy Recommendations

It is abundantly clear that Dhaka’s air pollution has serious detrimental effects on health and respiratory health. Furthermore, urgent and comprehensive action across multiple sectors is required:

Enhanced Emission Controls: Adopt and implement regulations for industrial emissions, brick kilns (with consideration of support for cleaner technologies), and vehicular emissions. Sustainable Urban Development: Improve public transport and construction and road dust management, increase green urban area, and waste management practices to eventually eliminate open burning (in Dhaka). Public Health Actions: Seek to release timely health advisories using the AQI forecasts to help target vulnerable populations with significant recommendations for less outdoor exposure during high pollution days. Prepare healthcare facilities for potential admissions related to pollution – the combination of pollution and COVID-19 is simply unprecedented. Research and Monitoring: Increase the number of air quality monitoring stations around Dhaka for better resolution; the health consequences of air pollution are continuing to be studied in the context of COVID-19, and further understanding is needed into the economic costs of air pollution. Evaluate the effectiveness of actions taken to date. Dealing with the combined threats of COVID-19 and air pollution: Acknowledge that poor air quality may contribute to worsened outcomes from respiratory infections, such as COVID-19. Public health actions should integrate efforts to address COVID-19 impacts and the situation with air pollution.

The National Air Quality Management Plan (2024–2030) is designed to reduce the PM2.5 concentrations established in this study; findings such as these are necessary evidence to motivate and implement the Plan beyond the current delays. [10]

## 5. Conclusion

In this comprehensive study covering five years (2020-2024), we have found a consistent, strong, and statistically significant relationship between daily elevated levels of ambient air pollution in Dhaka, and specific hospital admissions for asthma, COPD, acute bronchitis and pneumonia at Dhaka Medical College and Hospital. The elderly and young children are facing a disproportionate effect. Although the COVID-19 pandemic introduced some variability, the underlying threat of harmful air quality in Dhaka is an ongoing public health emergency that has not been rescinded due to the pandemic. Our findings highlight the need for immediate and ongoing action on the part of policymakers and stakeholders to deploy effective air pollution control initiatives as a means of reducing the burden of respiratory disease in Dhaka, and protecting the health of Dhaka’s public.

## Data Availability

All data produced in the present study are available upon reasonable request to the authors

## Acknowledgements

The authors express their gratitude to the administrative and medical records staff of Dhaka Medical College and Hospital for their invaluable assistance in facilitating data collection. We also thank the Department of Environment, Ministry of Environment, Forest and Climate Change, Bangladesh, for providing access to air quality monitoring data.

## Author Contributions

- **[Md Toukir Ahmed]:** Conceptualization, Methodology, Data Curation, Formal Analysis, Writing – Original Draft, Writing – Review & Editing.
- **[Md Rubel Hossain]:** Data Curation, Investigation, Software, Validation.

All authors have read and approved the final manuscript.

## Ethical Approval

Ethical approval for this study was obtained from the Institutional Review Board of Dhaka Medical College and Hospital (Ref: DMCH/IRB/2020/01) and the Bangladesh Medical Research Council (BMRC Ref: BMRC/NERC/2020-2024/237)

